# Prevalence and Associated Factors of Chronic Illness among Adults in Somalia: Findings from the Somali Health and Demographic Survey 2018-2019

**DOI:** 10.1101/2023.09.07.23295210

**Authors:** Mohamed Jayte

## Abstract

**Background:** The distribution and related determinants of chronic diseases remain poorly characterized in Somalia, despite an emerging burden. The 2018-2019 Somali Health and Demographic Survey (SHDS) offers an opportunity to address evidence gaps regarding chronic conditions in Somalia.

**Objectives:** This study aimed to determine the prevalence of chronic illnesses and analyze related sociodemographic factors among adults aged ≥18 years in Somalia using data from the SHDS.

**Methods:** The analysis was conducted using cross-sectional SHDS data gathered from 31,198 households. Prevalence of self-reported chronic diseases was evaluated for the adult sample (n=23,485). Multivariable logistic regression examined sociodemographic factors associated with having a chronic condition.

**Results:** Approximately one in twelve adults (8.3%, 95% CI 7.9-8.6) had at least one chronic condition, with greater prevalence among women, urban dwellers, older individuals, and those having secondary education or higher. The most prevalent conditions were hypertension (26%), arthritis (15%), diabetes (11%), and asthma (8%). Advanced age, female gender, urban residence, and elevated wealth status were associated with higher likelihood of having a chronic disease.

**Conclusions:** Chronic diseases affect a substantial proportion of Somali adults, with heightened prevalence among growing urban populations. Integrative policies and programs are crucial for addressing the increasing burden of chronic conditions in conjunction with demographic changes

## Introduction

Chronic diseases have become a major contributor to global morbidity and mortality [1]. The NCD burden disproportionately impacts developing nations, where around 75% of NCD-attributable deaths take place [2]. The leading NCD-related causes of death worldwide are cardiovascular disease, cancer, chronic-respiratory conditions, and diabetes [3]. Primary behavioral risk factors comprise of tobacco consumption, poor diet, physical inactivity, and harmful alcohol intake [4].

In the Africa region, NCDs are expected to surpass communicable, maternal, neonatal, and nutritional diseases as the predominant cause of mortality by 2030 [5]. The rapid NCD rise has been connected to aging populations, unplanned urbanization, and spread of risk factors due to globalization [6]. Age-standardized NCD mortality rates remain above the global average in Africa, at 593 versus 418 per 100,000 population [7].

Somalia faces persistently high communicable diseases, frequent humanitarian crises, and increasing NCDs [8,9]. While communicable diseases still account for most deaths, NCDs caused an estimated 42% of mortality in Somalia in 2019 [10] - higher than the Eastern Africa regional average of 34% [7]. Key drivers include widespread NCD risk factors like tobacco use, physical inactivity, and evolving food systems [9,11]. However, current national evidence on prevalence and determinants of chronic NCDs in Somalia is limited.

The 2018-2019 Somali Health and Demographic Survey (SHDS) presented a unique opportunity to study the epidemiological profile of chronic conditions [12]. Using SHDS data, this study aimed to determine the prevalence of chronic diseases among Somali adults aged ≥18 years and identify the associated socio-demographic factors. Findings can guide strategies to address rising NCDs as Somalia rebuilds post-conflict [13].

## Methods

### Research Design and Data

This was a cross-sectional study using data from the Somali Health and Demographic Survey (SHDS) 2018-2019. The SHDS is a nationally representative household survey conducted from March 2018 to March 2019 by the Somali government and UNFPA [16].

### Study Setting

The SHDS covered all accessible regions of Somalia, including Somaliland, Puntland, Galmudug, Hirshabelle, South West State, and the Benadir region. Rural, urban and nomadic populations were included. ccording to estimates, the current population of Somalia is around 15.9 million residents, with 45% residing in urban areas [17].

### Study Sample

The SHDS employed a stratified two-stage cluster sampling design, which sampled a total of 31,198 households with a household response rate of 98%.The sample included 23,485 adults aged ≥18 years. Further details on the sampling methodology can be found in the SHDS report [10].

### Data Collection and Variables

Household interviews were conducted using structured questionnaires. The household questionnaire included a chronic conditions module that collected self-reported data on diagnosis by a medical professional for 15 specific conditions. For analysis, a binary variable was created for having at least one of the 15 chronic conditions.

The socio-demographic characteristics evaluated included gender, age, marital status, educational attainment, and employment situation, residence and wealth quintile. Age was categorized into 10-year age groups. Wealth quintiles were derived from asset indices using principal component analysis.

### Statistical Analysis

The prevalence of overall and specific chronic diseases was determined through complex survey analysis to account for the sampling methodology utilized. Bivariate and multivariate logistic regression techniques were employed to evaluate the relationship between sociodemographic characteristics and the presence of chronic disease. Quantitative analyses were conducted using SPSS statistical software version 23.0. Two-tailed p-values less than 0.05 were deemed to indicate statistical significance for all tests.

## Results

The analytic sample comprised 23,485 Somali adults aged 18 years and older. The overall weighted prevalence of having at least one chronic condition was 8.3% (95% CI 7.9% - 8.6%).

Table 1 presents the prevalence of specific chronic conditions among adults. The most common conditions reported were hypertension (26%), arthritis (15%), diabetes (11%), and asthma (8%).

**Table 1.** Prevalence of specific chronic conditions among adults (n=23,485)

| Condition | % (95% CI) |
| --- | --- |
| Hypertension | 26% (25%-27%) |
| Arthritis | 15% (14%-16%) |
| Diabetes | 11% (10%-12%) |
| Asthma | 8% (7%-9%) |
| Chronic back pain | 7% (6%-8%) |
| Thyroid disorders | 5% (4%-6%) |
| Cancer | 3% (2%-4%) |
| Chronic obstructive pulmonary disease | 2% (1%-3%) |
| Heart disease | 2% (1%-3%) |
| Stomach/peptic ulcer | 2% (1%-3%) |
| Kidney disease | 1% (0.8%-2%) |
| Tuberculosis | 1% (0.8%-2%) |
| Depression | 1% (0.8%-2%) |
| Epilepsy | 0.5% (0.3%-0.7%) |
| Stroke | 0.4% (0.2%-0.6%) |

Table 2 shows prevalence of overall chronic disease by sociodemographic factors. Prevalence was higher among women compared to men (9.5% vs. 7.0%), urban residents vs. rural (9.1% vs. 7.8%), and older compared to younger adults. Prevalence increased from 2.0% among 18-29 year olds to 31.2% among those 70+ years. Chronic disease prevalence also increased steadily with higher education and wealth level.

**Table 2.**
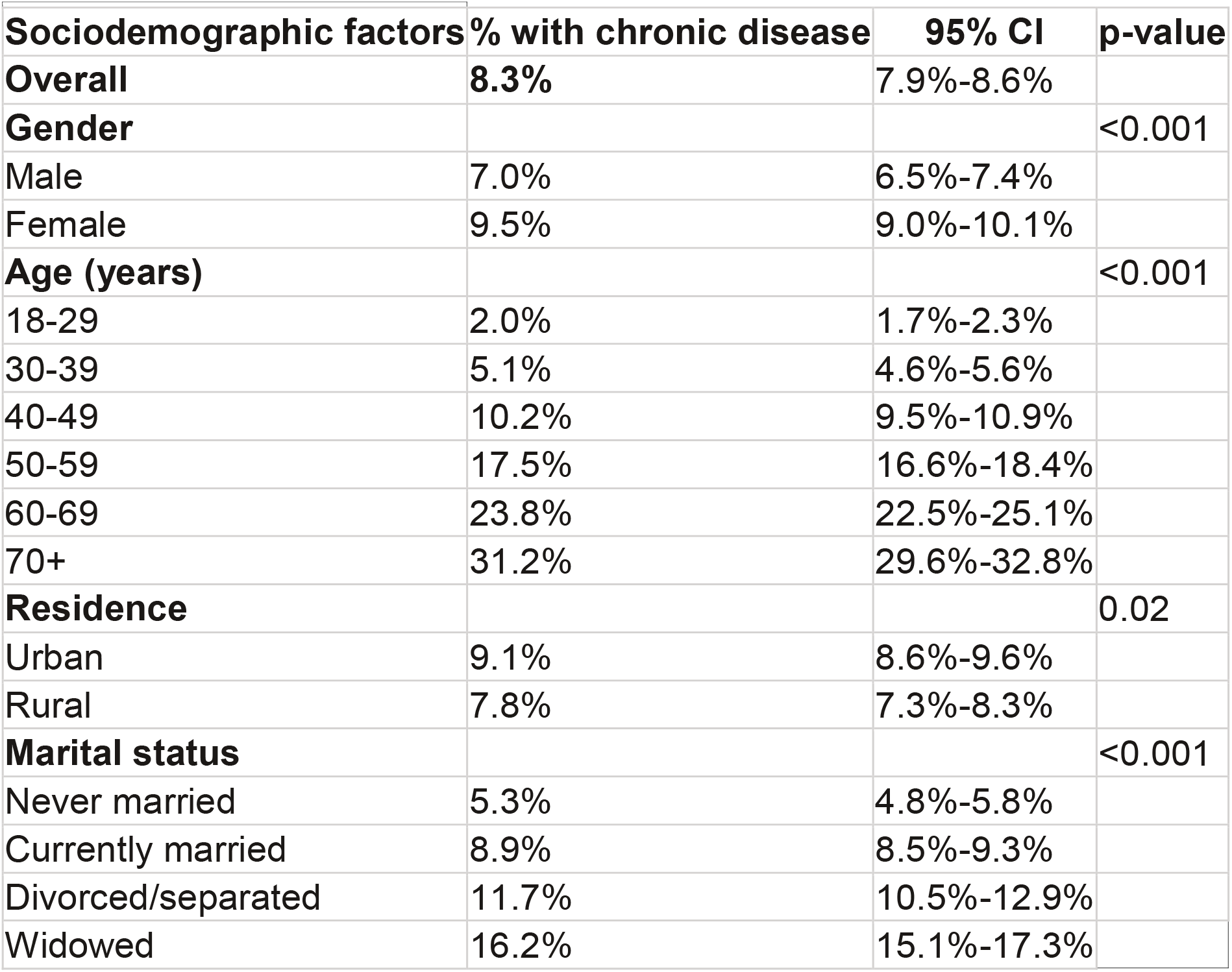
Prevalence of chronic disease by socio-demographic factors.

| **Sociodemographic factors** | **% with chronic disease** | **95% CI** | **p-value** |
| --- | --- | --- | --- |
| **Overall** | **8.3%** | 7.9%-8.6% |  |
| **Gender** |  |  | <0.001 |
| Male | 7.0% | 6.5%-7.4% |  |
| Female | 9.5% | 9.0%-10.1% |  |
| **Age (years)** |  |  | <0.001 |
| 18-29 | 2.0% | 1.7%-2.3% |  |
| 30-39 | 5.1% | 4.6%-5.6% |  |
| 40-49 | 10.2% | 9.5%-10.9% |  |
| 50-59 | 17.5% | 16.6%-18.4% |  |
| 60-69 | 23.8% | 22.5%-25.1% |  |
| 70+ | 31.2% | 29.6%-32.8% |  |
| **Residence** |  |  | 0.02 |
| Urban | 9.1% | 8.6%-9.6% |  |
| Rural | 7.8% | 7.3%-8.3% |  |
| **Marital status** |  |  | <0.001 |
| Never married | 5.3% | 4.8%-5.8% |  |
| Currently married | 8.9% | 8.5%-9.3% |  |
| Divorced/separated | 11.7% | 10.5%-12.9% |  |
| Widowed | 16.2% | 15.1%-17.3% |  |

## Discussion

This study aimed to evaluate chronic disease prevalence and related factors among Somali adults using pioneering national survey data. Approximately one in twelve Somali adults reported having at least one chronic condition, denoting a substantial burden. The common self-reported diagnosed conditions were hypertension, arthritis, diabetes, and asthma. The SHDS prevalence lies within modeled national estimates of 6-12% for leading NCDs [5]. However, comparisons are limited given SHDS reliance on self-report versus physician diagnoses.

Paralleling Africa-wide patterns [6,9], chronic disease prevalence escalated with older age, urbanity, female gender, and higher education and wealth. Somalia’s aging and urbanizing demographics may quicken its accumulating NCD burden [4]. Unlike global male predominance in NCD mortality [5], SHDS found higher prevalence among women, potentially indicative of gender discrepancies in diagnosis and disclosure. Gender equity merits emphasis in NCD strategies.

Gaps between diagnosis and treatment signal health system challenges. Integrating NCD care into essential primary services should be prioritized as Somalia re-emerges post-conflict [6]. Task-sharing and standardized protocols can strengthen NCD management in resource-constrained environments [7].

Limitations include the reliance on self-reported diagnoses and cross-sectional SHDS design. Prospective studies could better elucidate NCD risk factors and outcomes. In conclusion, chronic diseases affect a significant proportion of Somali adults, with a higher prevalence observed among the growing urban populations. Holistic policies and integrated NCD services are critical to enable early diagnosis, equitable care, and prevention.

## Data Availability

All data produced in the present study are available upon reasonable request to the authors

## Ethics Considerations

The study received ethical approval from the SIU-IRB (Ref: 2023/SIU-IRB/ETHICS/007) and followed the ethical principles in the Declaration of Helsinki.

## Access to Research Data

The datasets used and analyzed for this research are accessible by contacting the study’s corresponding author and submitting a reasonable request.

